# COVID-19 Vaccination Timing, Relative to Acute COVID-19, and Subsequent Risk of Long COVID

**DOI:** 10.1101/2025.04.22.25326224

**Authors:** Zachary Butzin-Dozier, Yunwen Ji, Lin-Chiun Wang, A. Jerrod Anzalone, Jeremy Coyle, Rachael V. Phillips, Rena C. Patel, Jing Sun, Eric Hurwitz, Sarang Deshpande, Junming (Seraphina) Shi, Andrew Mertens, Mark J. van der Laan, John M. Colford, Alan E. Hubbard, the National COVID Cohort Collaborative (N3C) Consortium

**Affiliations:** School of Public Health, University of California, Berkeley, Berkeley, CA USA; University of Nebraska Medical Center, Omaha, NE USA; University of Alabama at Birmingham, Birmingham, AL USA; Bloomberg School of Public Health, Johns Hopkins University, Baltimore, MD, USA; University of North Carolina at Chapel Hill, Chapel Hill, NC USA

## Abstract

**Objectives:** Long COVID is a debilitating condition that impacts millions of Americans, but patients and clinicians have little information on how to prevent this disorder. Vaccination is a vital tool in preventing acute COVID-19 and may confer additional protection against Long COVID. There is limited evidence regarding the optimal timing of COVID-19 vaccination (i.e., vaccination schedule) to minimize the risk of Long COVID.

**Methods:** We applied Longitudinal Targeted Maximum Likelihood Estimation to electronic health record (EHR) data from a retrospective cohort of patients vaccinated against COVID-19 between December 2021 and September 2022. We evaluated the association between binary COVID-19 vaccination status (two or more doses vs. zero doses) and 12-month Long COVID risk among patients diagnosed with acute COVID-19 between December 2021 and September 2022. In addition, we compared the 12-month cumulative risk of Long COVID (ICD-10 code U09.9) among patients diagnosed with acute COVID-19 one to three months after vaccination, three to five months after vaccination, or five to seven months after vaccination while adjusting for relevant high-dimensional baseline and time-dependent covariates.

**Results:** We analyzed EHR data from a retrospective cohort of 1,558,018 patients. In our binary cohort (*n* = 519,980), we found that vaccinated patients had a lower risk of Long COVID than unvaccinated patients (adjusted marginal risk ratio 0.84 (0.81, 0.88)). In our longitudinal cohort (*n* = 1,085,291), we did not find a significant difference in Long COVID risk comparing patients who were diagnosed with acute COVID-19 one to three months after vaccination versus patients who were diagnosed with COVID-19 three to five months (adjusted marginal risk ratio 0.93 (95% CI 0.62, 1.41) or 5 to 7 months (adjusted marginal risk ratio 1.06 (95% CI 0.72, 1.56)) after vaccination.

**Conclusions:** We found that COVID-19 vaccination before SARS-CoV-2 infection was protective against Long COVID, and we did not find that this protection significantly waned within 7 months after vaccination. These findings suggest that COVID-19 vaccination protects against Long COVID.

## INTRODUCTION

Researchers have made major steps toward understanding, preventing, and treating acute COVID-19, but Long COVID remains a debilitating and poorly understood condition. Five years have passed since the COVID-19 pandemic began, and many individuals have grown fatigued with repeated vaccination booster doses, particularly given the common perception of reduced severity of acute infection since the Omicron wave began in late 2021.^1–3^

Systematic reviews have noted a lack of randomized trials evaluating the relationship between COVID-19 vaccination and Long COVID and have observed considerable heterogeneity of observational findings of studies that address this research question.^4,5^ Brannock et al. found that COVID vaccination was protective against the subsequent risk of Long COVID using a large, nationally sampled US cohort (adjusted odds ratio 0.66, 95% CI (0.55, 0.80)).^6^ Although recent studies have found a protective effect of COVID vaccination on the risk of Long COVID,^6–8^ less is known about the relationship between the timing of vaccination and additional doses relative to acute COVID-19 and the development of Long COVID. Vaccination timing is important in designing and promoting effective clinical guidance for COVID-19 vaccination schedules and recommendations.

We sought to evaluate the hypothesis that greater time between the most recent vaccination and COVID-19 infection would be associated with a greater risk of Long COVID diagnosis. These findings may be relevant for policymakers, researchers, and clinicians. An improved understanding of the relationship between vaccination timing and Long COVID can inform policymakers’ guidelines regarding booster vaccination schedules, and these results can also be used by public health officials to motivate individuals to follow these recommendations.

## METHODS

Few studies have evaluated the relationship between vaccination timing and long-term sequelae of acute COVID, including Long COVID. Two significant barriers to this research question are (1) methodological challenges to causal inference in observational settings with longitudinal confounding, high dimensionality, and a large proportion of nonrandom missingness and (2) the lack of available longitudinal or biomarker data with sufficient sample size. These challenges can be overcome, respectively, through a targeted machine learning (i.e., causal inference) approach to analyzing data from the National Clinical Cohort Collaborative (N3C).

N3C includes electronic health records data for more than 22 million total patients, which are provided by 84 health institutions.^9^

We estimated our target causal parameter of interest while adjusting for relevant covariates and accounting for heterogeneous monitoring. We applied Longitudinal Targeted Maximum Likelihood Estimation to generate a marginal structural model (LTMLE, R package “ltmle”).^10–14^ LTMLE enables the estimation of causal parameters (in this case, hazard ratios) from nonparametric measures of association that are interpretable to non-statisticians (e.g., clinicians, policymakers), and the estimators used were doubly robust and maximally efficient. We estimated the parameters of interest using the LTMLE package and used generalized linear models as candidate algorithms to estimate the components of the longitudinal data-generating distribution. In our binary cohort, we applied Targeted Maximum Likelihood Estimation^15,16^ and defined binary exposure (yes/no) over the entire period of risk (see Binary cohort). See Supplemental Material 1 for details on our causal parameters of interest and relevant assumptions.

### Shared methods across longitudinal and binary cohorts

#### Exclusion criteria

As both documentation of COVID-19 vaccination and Long COVID diagnosis are highly heterogeneous by study site, we only included study sites that report at least 5% of their patients have at least one COVID-19 vaccination or booster dose and report that at least 1% of their patients have been diagnosed with Long COVID at some point during the N3C observation window (2018 to present). We excluded patients with a Long COVID diagnosis within the 4 weeks after the index COVID-19 infection date (as most definitions of Long COVID require that approximately one month must pass after acute COVID-19 for a patient to be considered “at risk” for Long COVID).^17–20^ *Covariates:* We included covariates that were associated with and plausibly caused (i.e., temporally preceded) the exposure and outcome (i.e., assessed at baseline).

Patient characteristics included sex, age (years), race, ethnicity, body mass index, immunocompromised status (based on documented diagnoses), systemic corticosteroid use, depression, chronic lung disease, hypertension, obesity, diabetes, tobacco use, asthma, date of inclusion (index vaccination date or acute COVID-19 date), and healthcare utilization rate (healthcare interactions per month; healthcare utilization is highly associated with Long COVID diagnosis).^21^ We adjusted for data partner and source common data model format to adjust for geospatial trends and systematic variations in health documentation formats.^21^ *Outcome*: Our primary outcome of interest was the cumulative incidence of Long COVID (ICD 10 code U09.9) 1 to 12 months after acute COVID-19.^17,18^

### Binary cohort: Vaccinated versus unvaccinated

We evaluated binary vaccination status (unvaccinated vs. vaccinated) before acute COVID-19. We hypothesized that COVID-19 vaccination before acute COVID-19 would be protective against Long COVID. *Inclusion criteria:* We included participants with documented COVID-19 between December 25, 2021 (CDC-reported data of Omicron variant dominance in the US)^22^ and September 25, 2022.^23^ *Exposure of interest:* We defined vaccination as a binary variable indicating whether the patient received at least two doses of a COVID-19 vaccination (or booster) before their index acute COVID-19 date. We excluded patients who only received one vaccination dose before acute COVID-19.

### Longitudinal cohort: Vaccination timing relative to acute COVID-19

We hypothesized that a greater time between the most recent vaccination and COVID-19 is associated with a greater risk of Long COVID diagnosis. *Inclusion criteria, sample size, and power:* We included participants with a documented COVID-19 vaccine or booster dose between December 25, 2021 (CDC-reported data of Omicron variant dominance in the US)^22^ and September 25, 2022 (to allow for sufficient follow-up time) (see Supplemental Material 2 for details on study timeline). *Exposures:* The primary exposure of interest was the time (months) between the initial vaccination or booster date and the date of acute COVID-19. We operationalized this exposure by defining “baseline” (start of follow-up) for each individual as the first COVID-19 vaccination or booster received between December 25, 2021, and September 25, 2022.^22^ We evaluated these patients’ acute COVID-19 status between 1 and 7 months following the vaccination date. We limited observation to this 6-month interval to account for the frequent missingness of vaccination data in N3C. By only including patients with a documented vaccination, at the time of vaccination, and observing subsequent vaccination and acute COVID-19 status for 7 months, we have created a cohort where we are confident in patient vaccination status during this observation period (as few patients received multiple vaccination doses during this observation period). During this interval, we also evaluated additional vaccination doses and healthcare utilization rate, as healthcare utilization is strongly associated with Long COVID diagnosis.^20,21^ If the patient was diagnosed with acute COVID-19 during this period, we monitored their Long COVID status in the 12 months following acute COVID-19. We partitioned time in two-month intervals, where each patient has a binary status for COVID-19 vaccination/booster, healthcare utilization (i.e., at least one healthcare interaction), and Long COVID diagnosis in each two-month interval (one to three months [1≤ *t* < 3], three to five months [3 ≤ *t* < 5], or five to seven [5 ≤ *t* < 7] months). *Analysis methods:* We applied Longitudinal Targeted Maximum Likelihood Estimation (LTMLE) to assess the causal impact of vaccination timing relative to COVID-19 on the risk of Long COVID^10–13^. These risk ratios reflect the relationship between time since vaccination and the risk of Long COVID.

#### Secondary analyses

Susceptibility to symptomatic COVID-19 soon after COVID-19 vaccination may indicate an underlying predisposition or risk for COVID-19, due to an immunocompromising condition. As a sensitivity analysis, we conducted a subgroup analysis of “low-risk” individuals to evaluate whether we observed similar temporal relationships between COVID-19 vaccination timing relative to acute COVID-19 and subsequent Long COVID risk.

This “low-risk” subgroup was defined by: age at acute COVID-19 between 18 and 65 years, no chronic lung disease, and no immunocompromised status.

**Table 1.**
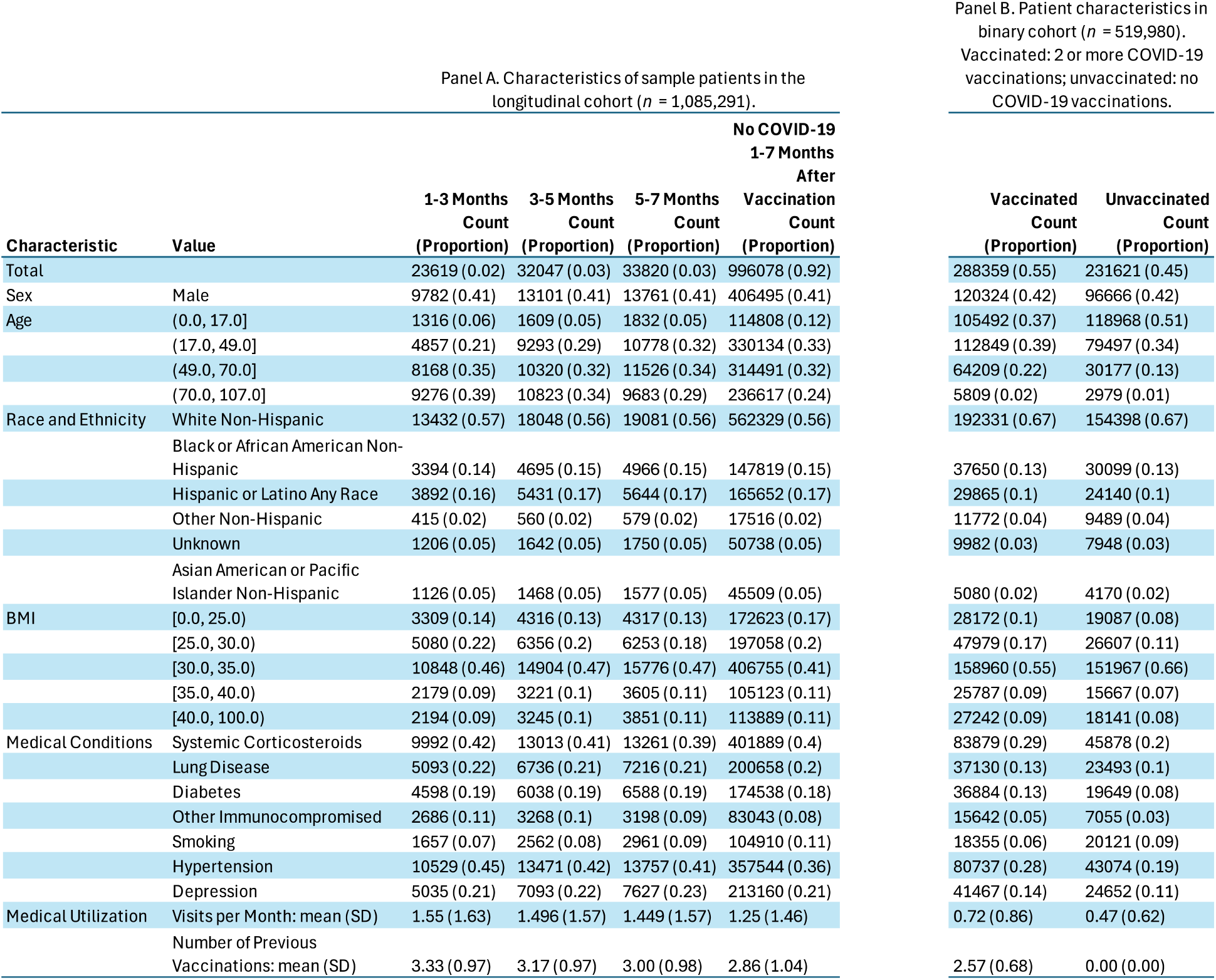
Characteristics of sample patients.

## RESULTS

Our binary cohort of vaccinated (2 or more doses) versus unvaccinated (0 doses) patients included a cohort of 670,452 patients (1,702,308 unique patients across longitudinal and binary cohorts), where 51% were vaccinated before acute COVID-19. Generally, vaccinated patients had higher healthcare utilization (0.7 versus 0.4 visits per month), a greater comorbidity burden (28% vs 17% systemic corticosteroid use, 13% vs. 9% lung disease, 12% vs. 6% diabetic, 5% vs. 2% immunocompromised, and 26% vs. 13% hypertensive) and were older (14% vs 6% over 70 years old) than unvaccinated patients.

Our longitudinal cohort included data from 1,085,291 patients who were vaccinated against COVID-19 between December 2021 and September 2022. Of these patients, 2% were diagnosed with acute COVID-19 one to three months after vaccination, 3% were diagnosed with acute COVID-19 three to five months after vaccination, 3% were diagnosed with acute COVID-19 5 to 7 months after vaccination, and 92% were not diagnosed with acute COVID-19 between 1 and 7 months after vaccination. Patients who were vaccinated 1 to 7 months before acute COVID-19 had similar healthcare utilization rates to one another (1.4 to 1.5 interactions per month), while patients without a documented case of acute COVID-19 during this range had a lower healthcare utilization rate (1.2 interactions per month). Patients who were vaccinated one to three months before acute COVID-19 had the most previous number of COVID-19 vaccinations (3.3), followed by 3-5 months (3.2), 5-7 months (3.0), and no acute COVID-19 within 1-7 months after vaccination (2.9). Patient age, body mass index, and comorbidity burden were similar across groups.

**Figure 1.**
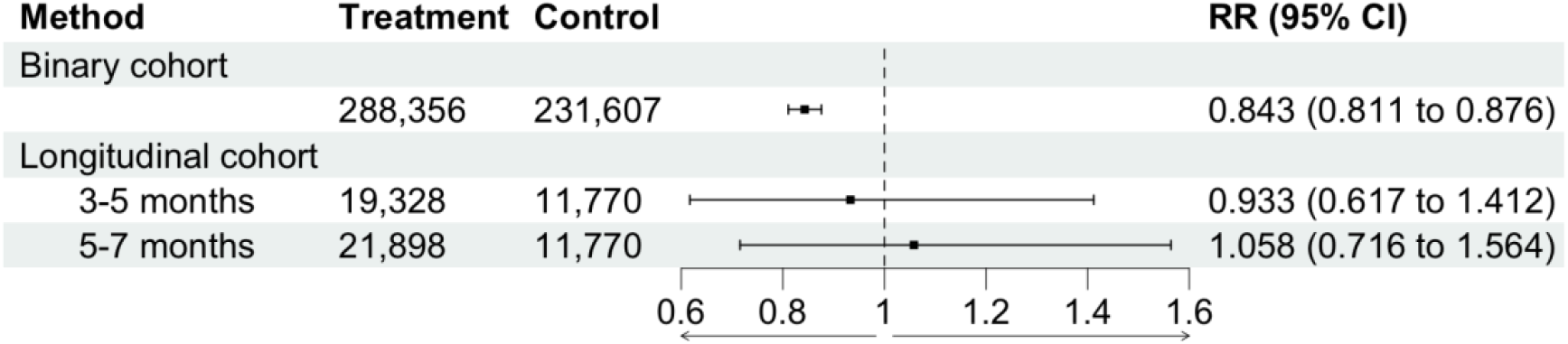
Relationship between vaccination status and 12-month cumulative incidence of Long COVID. In the binary cohort, “treatment” refers to patients with 2 or more COVID-19 vaccination doses, while “control” refers to patients with no COVID-19 vaccination doses. In the longitudinal cohort, “control” refers to patients who were vaccinated 1-3 months before acute COVID-19, while “treatment” refers to patients who were vaccinated either 3-5 months or 5-7 months before acute COVID-19, respectively. RR: Risk ratio.

### Binary cohort

In our adjusted model, we found that patients who were vaccinated against SARS-CoV-2 before acute COVID-19 had a lower risk of subsequent Long COVID, compared to patients with no documented SARS-CoV-2 vaccination before acute COVID-19 0.843 (0.811, 0.876), over the study period.

### Longitudinal cohort (vaccination timing and Long COVID)

We did not find a significant relationship between COVID-19 vaccination timing, relative to acute COVID-19, and subsequent risk of Long COVID in the first 7 months following vaccination. Compared to patients who experienced acute COVID-19 1-3 months after vaccination/booster, patients who experienced COVID-19 3-5 months after vaccination/booster had a Long COVID risk ratio of 0.933 (0.617, 1.412), and patients who experienced COVID-19 5-7 months after vaccination/booster had a Long COVID risk ratio of 1.058 (0.716, 1.564). The marginal adjusted, 12-month risk of Long COVID among patients who experienced acute COVID-19 one to three months after their most recent COVID-19 vaccination or booster was 0.005, for patients who experienced COVID-19 three to five months after vaccination/booster was 0.005, and for patients that experienced COVID-19 5 to 7 months after vaccination/booster was 0.006. We observed a similar relationship among low-risk patients (18 to 65 years old, not immunocompromised, and no lung disease) (Supplemental Material 3).

## DISCUSSION

We found that COVID-19 vaccination before acute COVID-19 was protective against subsequent Long COVID. We did not find evidence of waning protection of SARS-CoV-2 immunization concerning Long COVID in patients who experienced COVID-19 between 1 and 7 months following vaccination. These findings build on our understanding of COVID-19 vaccination effectiveness, particularly regarding Long COVID.

This study contributes to the body of literature demonstrating that vaccination against COVID-19 is protective against Long COVID.^4–6^ Our point estimate regarding the association between COVID-19 vaccination and subsequent Long COVID risk is more modest than some previous evaluations (e.g., risk ratio of 0.84 vs. odds ratio of 0.66)^6^ and is reflective of the heterogeneity in the documentation of both COVID-19 vaccination and Long COVID in electronic health records. These cumulative findings are consistent in demonstrating that SARS-CoV-2 vaccination before acute COVID-19 may prevent Long COVID.

Our findings regarding the relationship between vaccination timing and Long COVID are consistent with a previous retrospective cohort study that found that SARS-CoV-2 vaccination effectively protects against Long COVID for at least 6 months.^24^ Previous studies have shown that vaccination effectiveness against acute COVID-19 wanes over time.^25,26^ On the other hand, immunization against diseases like smallpox can lead to long-term protection even after circulating antibodies wane.^27^ Further research is needed to evaluate whether the effectiveness of SARS-CoV-2 vaccination, with respect to Long COVID, wanes over time.

Systematic reviews have noted a lack of randomized trials evaluating this relationship and have noted considerable heterogeneity in observational evaluations of this relationship, which supports the need for additional research in this area.^4,5^ In these settings, a causal inference (Targeted Machine Learning) approach can provide crucial evidence to approximate causal parameters via observational data.^10–12,16,28,29^ The approach described here provides a replicable method for evaluating the impact of post-acute COVID-19 vaccination timing on subsequent Long COVID risk, as several studies have hypothesized that SARS-COV-2 vaccination after acute COVID-19 may be protective against Long COVID.^30,31^. Furthermore, these methods can be replicated across a range of time-dependent, pre-exposure (e.g., pre-exposure prophylaxis), post-exposure (e.g., post-exposure prophylaxis), and sequential interventions to determine optimal intervention timing to minimize subsequent outcomes.

### Limitations

Due to the high variability of our findings (i.e., wide confidence intervals) in the longitudinal cohort, we do not interpret our results as a “true null” (i.e., there may be a relationship between vaccination timing and Long COVID). In other words, we do not conclude that the effectiveness of COVID-19 vaccination, with respect to Long COVID, does not wane over time. A study with greater power may detect a statistically significant relationship between vaccination timing and Long COVID. Future studies should further evaluate the relationship between vaccination timing and Long COVID through a data source with (1) higher quality vaccination data, (2) greater sensitivity of Long COVID outcome assessment, (3) a longer period of follow-up between vaccination and acute COVID-19, and (4) more complete patient health information (i.e., lower risk of bias due to differential healthcare utilization). In our longitudinal cohort, we addressed limitations in vaccination data quality (i.e., high missingness) by defining baseline as the date of a patient’s vaccination and limiting the exposure observation period to 7 months.

As is common with data from electronic health records (EHR), N3C has incomplete reporting of patient outcomes. This includes covariates related to baseline health and comorbidities, vaccination status, acute COVID-19 diagnosis, and Long COVID diagnosis. Previous studies using N3C have addressed these concerns.^6,32,33^ Notably, patient health information skews toward patients with high medical utilization rates, severe illness, or hospitalization.^32^ Therefore, our approach sought to characterize and account for this missingness, while transparently addressing limitations to generalizability. We did not evaluate effect modification by vaccination type (e.g., Pfizer vs. Johnson & Johnson) due to the high proportion of missingness of this information. One key method, in addition to longitudinal multivariate adjustment and weighting, is that we restricted study sites to only include data providers that reported at least 5% of their patients had at least one COVID-19 vaccination or booster dose and reported that at least 1% of their patients have been diagnosed with Long COVID at some point during the N3C observation window.

### Strengths

A major contribution of this study is the analytic method we applied. This study incorporated Longitudinal Targeted Maximum Likelihood Estimation to determine the relationship between high-dimensional covariates, multiple time-dependent exposures (vaccination and acute COVID-19), and longitudinal outcomes while intervening on heterogeneous monitoring. No single regression approach could characterize the exposure-outcome relationship due to time-dependent confounding. These methods can be applied to a range of research questions, particularly questions related to vaccination timing.

The data source, N3C, is another strength of this study. Given the potentially subtle association between vaccination timing and Long COVID, a large data source is needed to detect possible relationships. Furthermore, without randomization, high-dimensional covariate data is needed to adjust for imbalance. N3C provides this large and high-dimensional data source, which is necessary to address this research question.

## Conclusions

COVID-19 vaccination appears protective against Long COVID, and we did not find evidence that this protection wanes within 7 months of vaccination. This contributes to the body of literature supporting COVID-19 vaccination as a crucial public health measure.

## Data Availability

All analytic code and data are available in the N3C Enclave by request. Access to the N3C Data Enclave is managed by NCATS (https://ncats.nih.gov/research/research-activities/n3c/resources/data-access). Interested researchers must first complete a data use agreement, and next a data use request, in order to access the N3C Data Enclave. Once access is granted, the N3C data use committee must review and approve all use of data and the publication committee must approve all publications involving N3C data.

## DECLARATIONS

*Ethics approval and consent to participate:* This study was approved by the UC Berkeley Office for Protection of Human Subjects (2022-01-14980). The N3C data transfer to NCATS is performed under a Johns Hopkins University Reliance Protocol # IRB00249128 or individual site agreements with NIH. N3C received a waiver of consent from the NIH Institutional Review board and allows the secondary analysis of these data without additional consent.

## Consent to publish

The authors consent to the publication of this manuscript in its entirety.

## N3C Attribution

The analyses described in this manuscript were conducted with data or tools accessed through the NCATS N3C Data Enclave https://covid.cd2h.org and N3C Attribution & Publication Policy v 1.2-2020-08-25b supported by NCATS U24 TR002306, Axle Informatics Subcontract: NCATS-P00438-B, the Bill & Melinda Gates Foundation: OPP1165144, and the National Institutes of General Medical Sciences: U54GM115458 and 5U54GM104942-04. This research was possible because of the patients whose information is included within the data and the organizations (https://ncats.nih.gov/n3c/resources/data-contribution/data-transfer-agreement-signatories) and scientists who have contributed to the ongoing development of this community resource [https://doi.org/10.1093/jamia/ocaa196].

## Disclaimer

The N3C Publication committee confirmed that this manuscript (MSID: 2424.751) complies with N3C data use and attribution policies; however, the authors are solely responsible for its content, which does not necessarily represent the official views of the National Institutes of Health or the N3C program.

## IRB

The N3C data transfer to NCATS is performed under a Johns Hopkins University Reliance Protocol # IRB00249128 or individual site agreements with NIH. The N3C Data Enclave is managed under the authority of the NIH; information can be found at https://ncats.nih.gov/n3c/resources.

This research project was approved by the University of California, Berkeley Committee for the Protection of Human Subjects (CPHS protocol number 2022-01-14980). This approval is issued under University of California, Berkeley Federalwide Assurance #00006252.

## Individual Acknowledgements For Core Contributors

We gratefully acknowledge the following core contributors to N3C:

Adam B. Wilcox, Adam M. Lee, Alexis Graves, Alfred (Jerrod) Anzalone, Amin Manna, Amit Saha, Amy Olex, Andrea Zhou, Andrew E. Williams, Andrew Southerland, Andrew T. Girvin, Anita Walden, Anjali A. Sharathkumar, Benjamin Amor, Benjamin Bates, Brian Hendricks, Brijesh Patel, Caleb Alexander, Carolyn Bramante, Cavin Ward-Caviness, Charisse Madlock-Brown, Christine Suver, Christopher Chute, Christopher Dillon, Chunlei Wu, Clare Schmitt, Cliff Takemoto, Dan Housman, Davera Gabriel, David A. Eichmann, Diego Mazzotti, Don Brown, Eilis Boudreau, Elaine Hill, Elizabeth Zampino, Emily Carlson Marti, Emily R. Pfaff, Evan French, Farrukh M Koraishy, Federico Mariona, Fred Prior, George Sokos, Greg Martin, Harold Lehmann, Heidi Spratt, Hemalkumar Mehta, Hongfang Liu, Hythem Sidky, J.W. Awori Hayanga, Jami Pincavitch, Jaylyn Clark, Jeremy Richard Harper, Jessica Islam, Jin Ge, Joel Gagnier, Joel H. Saltz, Joel Saltz, Johanna Loomba, John Buse, Jomol Mathew, Joni L. Rutter, Julie A.

McMurry, Justin Guinney, Justin Starren, Karen Crowley, Katie Rebecca Bradwell, Kellie M. Walters, Ken Wilkins, Kenneth R. Gersing, Kenrick Dwain Cato, Kimberly Murray, Kristin Kostka, Lavance Northington, Lee Allan Pyles, Leonie Misquitta, Lesley Cottrell, Lili Portilla, Mariam Deacy, Mark M. Bissell, Marshall Clark, Mary Emmett, Mary Morrison Saltz, Matvey B. Palchuk, Melissa A. Haendel, Meredith Adams, Meredith Temple-O’Connor, Michael G. Kurilla, Michele Morris, Nabeel Qureshi, Nasia Safdar, Nicole Garbarini, Noha Sharafeldin, Ofer Sadan, Patricia A. Francis, Penny Wung Burgoon, Peter Robinson, Philip R.O. Payne, Rafael Fuentes, Randeep Jawa, Rebecca Erwin-Cohen, Rena Patel, Richard A. Moffitt, Richard L. Zhu, Rishi Kamaleswaran, Robert Hurley, Robert T. Miller, Saiju Pyarajan, Sam G. Michael, Samuel Bozzette, Sandeep Mallipattu, Satyanarayana Vedula, Scott Chapman, Shawn T. O’Neil, Soko Setoguchi, Stephanie S. Hong, Steve Johnson, Tellen D. Bennett, Tiffany Callahan, Umit Topaloglu, Usman Sheikh, Valery Gordon, Vignesh Subbian, Warren A. Kibbe, Wenndy Hernandez, Will Beasley, Will Cooper, William Hillegass, Xiaohan Tanner Zhang. Details of contributions available at covid.cd2h.org/core-contributors

## Data Partners with Released Data

The following institutions whose data is released or pending:

Available: Advocate Health Care Network — UL1TR002389: The Institute for Translational Medicine (ITM) • Boston University Medical Campus — UL1TR001430: Boston University Clinical and Translational Science Institute • Brown University — U54GM115677: Advance Clinical Translational Research (Advance-CTR) • Carilion Clinic — UL1TR003015: iTHRIV Integrated Translational health Research Institute of Virginia • Charleston Area Medical Center — U54GM104942: West Virginia Clinical and Translational Science Institute (WVCTSI) • Children’s Hospital Colorado — UL1TR002535: Colorado Clinical and Translational Sciences Institute • Columbia University Irving Medical Center — UL1TR001873: Irving Institute for Clinical and Translational Research • Duke University — UL1TR002553: Duke Clinical and Translational Science Institute • George Washington Children’s Research Institute — UL1TR001876: Clinical and Translational Science Institute at Children’s National (CTSA-CN) • George Washington University — UL1TR001876: Clinical and Translational Science Institute at Children’s National (CTSA-CN) • Indiana University School of Medicine — UL1TR002529: Indiana Clinical and Translational Science Institute • Johns Hopkins University — UL1TR003098: Johns Hopkins Institute for Clinical and Translational Research • Loyola Medicine — Loyola University Medical Center • Loyola University Medical Center — UL1TR002389: The Institute for Translational Medicine (ITM) • Maine Medical Center — U54GM115516: Northern New England Clinical & Translational Research (NNE-CTR) Network • Massachusetts General Brigham — UL1TR002541: Harvard Catalyst • Mayo Clinic Rochester — UL1TR002377: Mayo Clinic Center for Clinical and Translational Science (CCaTS) • Medical University of South Carolina — UL1TR001450: South Carolina Clinical & Translational Research Institute (SCTR) • Montefiore Medical Center — UL1TR002556: Institute for Clinical and Translational Research at Einstein and Montefiore • Nemours — U54GM104941: Delaware CTR ACCEL Program • NorthShore University HealthSystem — UL1TR002389: The Institute for Translational Medicine (ITM) • Northwestern University at Chicago — UL1TR001422: Northwestern University Clinical and Translational Science Institute (NUCATS) • OCHIN — INV-018455: Bill and Melinda Gates Foundation grant to Sage Bionetworks • Oregon Health & Science University — UL1TR002369: Oregon Clinical and Translational Research Institute • Penn State Health Milton S. Hershey Medical Center — UL1TR002014: Penn State Clinical and Translational Science Institute • Rush University Medical Center — UL1TR002389: The Institute for Translational Medicine (ITM) • Rutgers, The State University of New Jersey — UL1TR003017: New Jersey Alliance for Clinical and Translational Science • Stony Brook University — U24TR002306 • The Ohio State University — UL1TR002733: Center for Clinical and Translational Science • The State University of New York at Buffalo — UL1TR001412: Clinical and Translational Science Institute • The University of Chicago — UL1TR002389: The Institute for Translational Medicine (ITM) • The University of Iowa — UL1TR002537: Institute for Clinical and Translational Science • The University of Miami Leonard M. Miller School of Medicine — UL1TR002736: University of Miami Clinical and Translational Science Institute • The University of Michigan at Ann Arbor — UL1TR002240: Michigan Institute for Clinical and Health Research • The University of Texas Health Science Center at Houston — UL1TR003167: Center for Clinical and Translational Sciences (CCTS) • The University of Texas Medical Branch at Galveston — UL1TR001439: The Institute for Translational Sciences • The University of Utah — UL1TR002538: Uhealth Center for Clinical and Translational Science • Tufts Medical Center — UL1TR002544: Tufts Clinical and Translational Science Institute • Tulane University — UL1TR003096: Center for Clinical and Translational Science • University Medical Center New Orleans — U54GM104940: Louisiana Clinical and Translational Science (LA CaTS) Center • University of Alabama at Birmingham — UL1TR003096: Center for Clinical and Translational Science • University of Arkansas for Medical Sciences — UL1TR003107: UAMS Translational Research Institute • University of Cincinnati — UL1TR001425: Center for Clinical and Translational Science and Training • University of Colorado Denver, Anschutz Medical Campus — UL1TR002535: Colorado Clinical and Translational Sciences Institute • University of Illinois at Chicago — UL1TR002003: UIC Center for Clinical and Translational Science • University of Kansas Medical Center — UL1TR002366: Frontiers: University of Kansas Clinical and Translational Science Institute • University of Kentucky — UL1TR001998: UK Center for Clinical and Translational Science • University of Massachusetts Medical School Worcester — UL1TR001453: The UMass Center for Clinical and Translational Science (UMCCTS) • University of Minnesota — UL1TR002494: Clinical and Translational Science Institute • University of Mississippi Medical Center — U54GM115428: Mississippi Center for Clinical and Translational Research (CCTR) • University of Nebraska Medical Center — U54GM115458: Great Plains IDeA-Clinical & Translational Research • University of North Carolina at Chapel Hill — UL1TR002489: North Carolina Translational and Clinical Science Institute • University of Oklahoma Health Sciences Center — U54GM104938: Oklahoma Clinical and Translational Science Institute (OCTSI) • University of Rochester — UL1TR002001: UR Clinical & Translational Science Institute • University of Southern California — UL1TR001855: The Southern California Clinical and Translational Science Institute (SC CTSI) • University of Vermont — U54GM115516: Northern New England Clinical & Translational Research (NNE-CTR) Network • University of Virginia — UL1TR003015: iTHRIV Integrated Translational health Research Institute of Virginia • University of Washington — UL1TR002319: Institute of Translational Health Sciences • University of Wisconsin-Madison — UL1TR002373: UW Institute for Clinical and Translational Research • Vanderbilt University Medical Center — UL1TR002243: Vanderbilt Institute for Clinical and Translational Research • Virginia Commonwealth University — UL1TR002649: C. Kenneth and Dianne Wright Center for Clinical and Translational Research • Wake Forest University Health Sciences — UL1TR001420: Wake Forest Clinical and Translational Science Institute • Washington University in St. Louis — UL1TR002345: Institute of Clinical and Translational Sciences • Weill Medical College of Cornell University — UL1TR002384: Weill Cornell Medicine Clinical and Translational Science Center • West Virginia University — U54GM104942: West Virginia Clinical and Translational Science Institute (WVCTSI)

Submitted: Icahn School of Medicine at Mount Sinai — UL1TR001433: ConduITS Institute for Translational Sciences • The University of Texas Health Science Center at Tyler — UL1TR003167: Center for Clinical and Translational Sciences (CCTS) • University of California, Davis — UL1TR001860: UCDavis Health Clinical and Translational Science Center • University of California, Irvine — UL1TR001414: The UC Irvine Institute for Clinical and Translational Science (ICTS) • University of California, Los Angeles — UL1TR001881: UCLA Clinical Translational Science Institute • University of California, San Diego — UL1TR001442: Altman Clinical and Translational Research Institute • University of California, San Francisco — UL1TR001872: UCSF Clinical and Translational Science Institute

Pending: Arkansas Children’s Hospital — UL1TR003107: UAMS Translational Research

Institute • Baylor College of Medicine — None (Voluntary) • Children’s Hospital of Philadelphia — UL1TR001878: Institute for Translational Medicine and Therapeutics • Cincinnati Children’s Hospital Medical Center — UL1TR001425: Center for Clinical and Translational Science and Training • Emory University — UL1TR002378: Georgia Clinical and Translational Science Alliance • HonorHealth — None (Voluntary) • Loyola University Chicago — UL1TR002389: The Institute for Translational Medicine (ITM) • Medical College of Wisconsin — UL1TR001436: Clinical and Translational Science Institute of Southeast Wisconsin • MedStar Health Research Institute — UL1TR001409: The Georgetown-Howard Universities Center for Clinical and Translational Science (GHUCCTS) • MetroHealth — None (Voluntary) • Montana State University — U54GM115371: American Indian/Alaska Native CTR • NYU Langone Medical Center — UL1TR001445: Langone Health’s Clinical and Translational Science Institute • Ochsner Medical Center — U54GM104940: Louisiana Clinical and Translational Science (LA CaTS) Center • Regenstrief Institute — UL1TR002529: Indiana Clinical and Translational Science Institute • Sanford Research — None (Voluntary) • Stanford University — UL1TR003142: Spectrum: The Stanford Center for Clinical and Translational Research and Education • The Rockefeller University — UL1TR001866: Center for Clinical and Translational Science • The Scripps Research Institute — UL1TR002550: Scripps Research Translational Institute • University of Florida — UL1TR001427: UF Clinical and Translational Science Institute — University of New Mexico Health Sciences Center — UL1TR001449: University of New Mexico Clinical and Translational Science Center • University of Texas Health Science Center at San Antonio — UL1TR002645: Institute for Integration of Medicine and Science • Yale New Haven Hospital — UL1TR001863: Yale Center for Clinical Investigation

## Authors statement

Authorship was determined using ICMJE recommendations.

ZB: Generated research question, drafted manuscript, managed project timeline, and coordinated analysis.

YJ, LW, JA, JC, RVP, RCP, JS, SD, EH, JS, AM, ML, JC, and AH: Provided oversight on study design and analysis plan, supported analysis, reviewed manuscript, provided feedback, and

*Inclusion and ethics statement:* All co-authors and collaborators included in this manuscript have fulfilled the criteria for authorship.

## REFERENCES

1. Shah, A. & Coiado, O. C. COVID-19 vaccine and booster hesitation around the world: A literature review. Front Med (Lausanne) 9, 1054557 (2022).

2. Rzymski, P. & Szuster-Ciesielska, A. The COVID-19 Vaccination Still Matters: Omicron Variant Is a Final Wake-Up Call for the Rich to Help the Poor. Vaccines (Basel) 10, (2022).

3. Huang, M. et al. COVID-19 vaccine booster dose hesitancy among key groups: A cross-sectional study. Hum Vaccin Immunother 19, 2166323 (2023).

4. Chow, N. K. N. et al. The effect of pre-COVID and post-COVID vaccination on long COVID: A systematic review and meta-analysis. J Infect 89, 106358 (2024).

5. Byambasuren, O., Stehlik, P., Clark, J., Alcorn, K. & Glasziou, P. Effect of covid-19 vaccination on long covid: systematic review. BMJ Med 2, e000385 (2023).

6. Brannock, M. D. et al. Long COVID risk and pre-COVID vaccination in an EHR-based cohort study from the RECOVER program. Nature Communications 14, 2914 (2023).

7. Notarte, K. I. et al. Impact of COVID-19 vaccination on the risk of developing long-COVID and on existing long-COVID symptoms: A systematic review. eClinicalMedicine 53, (2022).

8. Azzolini, E. et al. Association Between BNT162b2 Vaccination and Long COVID After Infections Not Requiring Hospitalization in Health Care Workers. JAMA 328, 676–678 (2022).

9. National Institutes of Health. About the National COVID Cohort Collaborative. National Center for Advancing Translational Sciences https://ncats.nih.gov/n3c/about (2023).

10. van der Laan, M. J. & Rose, S. Targeted Learning: Causal Inference for Observational and Experimental Data. (Springer, New York, 2011).

11. van der Laan, M. J. & Rose, S. Targeted Learning in Data Science: Causal Inference for Complex Longitudinal Studies. (Springer Berlin Heidelberg, New York, NY, 2017).

12. van der Laan, M. et al. Targeted Learning in R: Causal Data Science with the Tlverse Software Ecosystem. (2023).

13. Lendle, S. D., Schwab, J., Petersen, M. L. & van der Laan, M. J. ltmle: An R Package Implementing Targeted Minimum Loss-Based Estimation for Longitudinal Data. J. Stat. Soft. 81, 1–21 (2017).

14. Rytgaard, H. C. W., Eriksson, F. & van der Laan, M. Estimation of time-specific intervention effects on continuously distributed time-to-event outcomes by targeted maximum likelihood estimation. arXiv preprint 2106.11009 (2021).

15. Schuler, M. S. & Rose, S. Targeted Maximum Likelihood Estimation for Causal Inference in Observational Studies. Am J Epidemiol 185, 65–73 (2017).

16. Coyle, J. R. et al. Targeted Learning. in Wiley StatsRef: Statistics Reference Online 1–20 (2023). doi:10.1002/9781118445112.stat08414.

17. Pfaff, E. R. et al. Coding long COVID: characterizing a new disease through an ICD-10 lens. BMC Med 21, 58 (2023).

18. ICD10 Data. 2023 ICD-10-CM Diagnosis Code U09.9. ICD10data.com https://www.icd10data.com/ICD10CM/Codes/U00-U85/U00-U49/U09-/U09.9 (2023).

19. McGrath, L. J. et al. Use of the Postacute Sequelae of COVID-19 Diagnosis Code in Routine Clinical Practice in the US. JAMA Netw Open 5, e2235089 (2022).

20. Butzin-Dozier, Z. et al. SSRI use during acute COVID-19 and risk of Long COVID among patients with depression. BMC Medicine 22, 445 (2024).

21. Butzin-Dozier, Z. et al. Predicting Long COVID in the National COVID Cohort Collaborative Using Super Learner: Cohort Study. JMIR Public Health Surveill 10, e53322 (2024).

22. Centers for Disease Control and Prevention. COVID data tracker: Monitoring variant proportions. https://covid.cdc.gov/covid-data-tracker/#variant-proportions (2023).

23. Pfaff, E., Kostka, K., Miller, R. & Beasley, W. COVID-19 Phenotype Documentation, Version 4.0. N3C Phenotype & Data Acquisition Workstream. Github.

24. Razzaghi, H. et al. Vaccine Effectiveness Against Long COVID in Children. Pediatrics 153, (2024).

25. Chemaitelly, H. & Abu-Raddad, L. J. Waning effectiveness of COVID-19 vaccines. Lancet 399, 771–773 (2022).

26. Menegale, F. et al. Evaluation of Waning of SARS-CoV-2 Vaccine-Induced Immunity: A Systematic Review and Meta-analysis. JAMA Netw Open 6, e2310650 (2023).

27. Akter, F. et al. Effect of prior immunisation with smallpox vaccine for protection against human Mpox: A systematic review. Rev Med Virol 33, e2444 (2023).

28. Pearl, J. Causality: Models, Reasoning, and Inference. (Cambridge University Press, Cambridge, U.K.; New York, 2000).

29. Pearl, J., Glymour, M. & Jewell, N. P. Causal Inference in Statistics: A Primer. (Wiley, Chichester, West Sussex, 2016).

30. Quach, T. C. et al. Post-COVID-19 Vaccination and Long COVID: Insights from Patient-Reported Data. Vaccines (Basel) 12, (2024).

31. Chow, N. K. N. et al. The effect of pre-COVID and post-COVID vaccination on long COVID: A systematic review and meta-analysis. J Infect 89, 106358 (2024).

32. Pfaff, E. R. et al. Identifying who has long COVID in the USA: a machine learning approach using N3C data. Lancet Digit Health 4, e532–e541 (2022).

33. Sun, J. et al. Association Between Immune Dysfunction and COVID-19 Breakthrough Infection After SARS-CoV-2 Vaccination in the US. JAMA Intern Med 182, 153–162 (2022).

