## Supplemental Material 1 for "COVID-19 Vaccination Timing, Relative to Acute COVID-19, and Subsequent Risk of Long COVID"

April 2025

#### 1 Longitudinal analysis

We observe $n$ independent and identically distributed samples of

$$O = (L(0), C(1), L(1), A(1), ..., C(K), L(K), A(K), Y = L(K+1)) \sim P_0$$

where each $k \in \{1, ..., K\}$ corresponds to a two-month interval. Here, $L(0)$ contains baseline covariates such as age and sex, and $L(k)$ for $k \geq 1$ includes time-varying covariates (e.g., vaccinations and visit indicators). The treatment variables $A(k)$ represent the incidence of COVID-19 infection at time $k$, while $C(k)$ indicates censoring (death). The outcome $Y$ is the incidence of long COVID, measured at the final time point.

Following the notations in the LTMLE literature [1], we use an overbar with a time index to denote the entire history of a variable up to that point. For example, $\bar{L}(k)$ represents $(L(0), L(1), ..., L(k))$. If the time index is omitted, the history is taken up to the final time, so $\bar{L} = \bar{L}(K+1)$ and $\bar{A} = \bar{A}(K+1)$. We consider a static treatment regime $a = (a_k : k \in \{0, \ldots, K+1\})$.

Our target parameter is then defined as the causal risk ratio of $a$ relative to a reference regime $a^{ref}$:

$$RR = \frac{E_{P_0}(Y^a)}{E_{P_0}(Y^{a^{ref}})},$$

where $a^{ref} = (1, 0, ..., 0)$ specifies COVID incidence in the first 1–3 months after baseline, and $a$ might be $(0, 1, 0, ..., 0)$ for incidence in months 3–5 or $(0, 0, 1, 0, ..., 0)$ for months 5–7.

To identify this causal parameter, we assume:

- Sequential randomization assumption: $Y^a \perp A_t \mid pa(A_t)$ for $t = 1, \ldots, T$
- Positivity assumption: $P_0\left(A(k) = a_k \mid \bar{L}(k), \bar{A}(k-1) = a_{k-1}\right) > 0$ almost everywhere.

Under these assumptions , the marginal causal risk can be identified via the longitudinal G-computation formula [2].

#### 2 Binary analysis

We observe $n$ independent and identically distributed copies of $O$ with a data structure $O = (W, A, \Delta, \Delta Y) \sim P_0 \in \mathcal{M}$, where $W \in R^p$ denotes baseline covariates, $A \in \{0, 1\}$ is the indicator of complete vaccination, and $\Delta Y \in \{0, 1\}$ is observed Long COVID diagnosis. $\Delta$ refers to patient observation during the outcome period. The statistical model for the probability distribution $P_0$ is denoted by $\mathcal{M}$.

Our target parameter is

$$\frac{EY^1}{EY^0},$$

where $Y^a$ is the counterfactual result under treatment $A = a$. Under standard positivity and no unmeasured confounding assumptions, the target parameter can be identified by the following statistical parameter:

$$E_{P_0}(E_{P_0}(Y \mid A = 1, W, \Delta = 1) - E_{P_0}(Y \mid A = 0, W, \Delta = 1))$$

### References

[1] Samuel D. Lendle et al. "ltmle: An R Package Implementing Targeted Minimum Loss-Based Estimation for Longitudinal Data". In: *Journal of Statistical Software* 81.1 (2017), pp. 1–21. DOI: 10.18637/jss.v081.i01. URL: https://www.jstatsoft.org/index.php/jss/article/view/v081i01.

[2] James Robins. "A new approach to causal inference in mortality studies with a sustained exposure period—application to control of the healthy worker survivor effect". In: *Mathematical Modelling* 7.9 (1986), pp. 1393–1512. ISSN: 0270-0255. DOI: https://doi.org/10.1016/0270-0255(86)90088-6. URL: https://www.sciencedirect.com/science/article/pii/0270025586900886.
