## Supplemental Material 2 for "COVID-19 Vaccination Timing, Relative to Acute COVID-19, and Subsequent Risk of Long COVID"

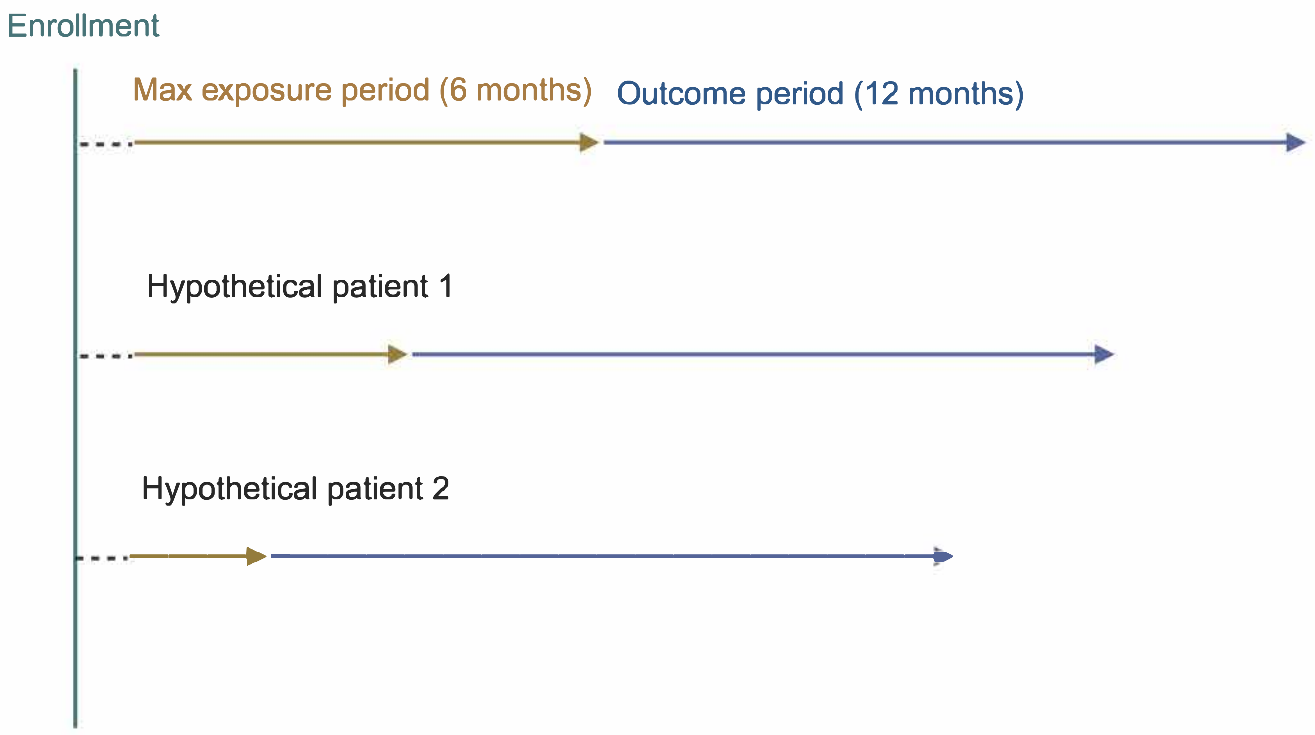


Enrollment: initial COVID-19 vaccination between December 25, 2021 and September 25, 2022. We adjusted for baseline patient variables at this point (see Methods).

Exposure period: we assessed patient acute COVID-19 status months 1-7 after enrollment. We excluded patients with COVID-19 diagnoses within 1 month of vaccination. We discretized over 2 month intervals and assessed healthcare utilization and additional vaccination doses in each interval

Outcome period: We assessed patient Long COVID status in the 12 months following acute COVID-19. The outcome period began if/when the patient was diagnosed with acute COVID-19. We discretized over 2 months intervals and assessed healthcare utilization and additional vaccination doses in each interval.
